# Features of α-Hydroxybutyrate Dehydrogenase during various specific periods in COVID-19 patients within Xiangyang, China: a cohort study

**DOI:** 10.1101/2020.10.28.20221127

**Authors:** Haoming Zhu, Gaojing Qu, Hui Yu, Guoxin Huang, Lei Chen, Meiling Zhang, Shanshan Wan, Bin Pei

**Affiliations:** Center of Evidence-Based Medicine, Xiangyang No.1 People’s Hospital, Hubei University of Medicine, Xiangyang, 441000, China; Postgraduate Training Basement of Jinzhou Medical University, Xiangyang No.1 People’s Hospital, Hubei University of Medicine, Xiangyang, 441000, China

**Keywords:** Coronavirus disease-2019, respiratory infection, clinical features, laboratory findings

## Abstract

**Background:** Coronavirus disease-2019 (COVID-19) has spread all over the world and brought extremely huge losses. There is no study to systematically analyse the features of hydroxybutyrate dehydrogenase (α-HBDH) in COVID-19 patients during the periods before and after illness progression, before death and course from exposure onset.

**Methods:** We collected all included patients’ general information, clinical type, α-HBDH value and outcome, and analyzed α-HBDH values within different initial time and different periods.

**Results:** In the first 30 days after symptom onset, the α-HBDH median value was 156.33 U/L. The first test of α-HBDH since exposure onset appeared on the 8th day, it increased from the 8th day to 18th day and decreased after the 18th day. α-HBDH median value showed a slight change until it started to increase 1 day before transforming to severe type, while it continued to increase during 4 days before and after transforming to critical type. The α-HBDH median value ranged from 191.11 U/L to 455.11U/L before death.

**Conclusions:** α-HBDH value increases in some COVID-19 patients, obviously in severe type, critical type and death patients, and mainly in 18 days after exposure onset and 10 days after symptom onset. α-HBDH increases 1 day before transforming to severe type, continues to increase in critical type and death patients, increases rapidly 5 days before death. The increase of α-HBDH suggests that COVID-19 patients have tissues and organs damage, mainly in heart. In brief, α-HBDH is an important indicator to judge the severity and prognosis of COVID-19.

## 1 Introduction

COVID-19 has spread around the world since the outbreak in December 2019, which is caused by SARS-Cov-2, and has attracted the attention of the global because of its high transmission ability, morbidity and mortality^[1-4]^. On January 30, the World Health Organization (WHO) identified COVID-19 as a public health emergency of international concern^[5]^. As of August 31, 2020, a cumulative total of nearly 25 million cases and 800,000 deaths have been reported ^[6]^.

Lactate dehydrogenase (LDH) is one of the important enzymes in glycolysis and gluconeogenesis. It mainly catalyzes the transformation between lactic acid and pyruvate. Its enzymatic reaction is: pyruvate + NADH+H^+^ ⇌ lactic acid + NAD^+^. LDH consists of five isozymes composed of different combinations of H and M subunits: LDH1 (H4), LDH2 (H3M), LDH3 (H2M2), LDH4 (HM3) and LDH5 (M4). α-HBDH is tested by the α-ketoacid, a substrate, to determine the LDH activity. Additionally, the activity of LDH1 and LDH2 with more H subunits is described by α-HBDH activity because of the high affinity for this substrate to the H subunit in LDH. α-HBDH mainly distributes in the heart, brain, kidney and red blood cells, and the activity of the enzyme in the heart is more than half of the total enzyme activity, and it level increases in the progression of cor pulmonale, leukemia and tumor. Moreover, the extent of the increase is closely related to the tissue and organ injury, which can be used as an auxiliary diagnostic index in clinical practice^[7-11]^.

Previous studies have shown that α-HBDH is related to COVID-19. Cen Y et al. observed 1007 mild and moderate COVID-19 patients for 28 days. It was found that the higher the α-HBDH value, the greater the risk of progression to severe or critical type^[12]^. Compared with other pneumonia types, the α-HBDH level in COVID-19 patients was significantly higher, and the α-HBDH value of severe group was higher than that of non-severe group^[13,14]^. When complicated with cardiovascular disease or gastrointestinal symptoms, the α-HBDH in COVID-19 patients was much higher as well^[15,16]^. However, there are no reports to study the α-HBDH features in COVID-19 patients during the period from symptom onset, from exposure onset, before and after illness progression and before death. In this study, we analyzed the changes of α-HBDH values in COVID-19 patients during these specific period.

## 2 Materials and Methods

### 2.1 Study Design

This study was a bidirectional observational cohort study. It was established on Feb 9, 2020, all suspected and confirmed COVID-19 patients hospitalized in Xiangyang No.1 People’s Hospital before Feb 28, 2020 were included in this cohort. All information was traced back to Jan 23, 2020. The last day of follow-up was Mar 28, 2020. Admission standard and clinical classifications were made according to the Diagnosis Guidance for Novel Coronavirus Pneumonia ^[17]^. The study was approved by the ethics review board of Xiangyang No.1 People’s Hospital (No. 2020GCP012) and registered at the Chinese Clinical Trial Registry as ChiCTR2000031088. Informed consent from patients has been exempted since this study does not involve patients’ personal privacy.

### 2.2 Data Collection

The pre-designed extraction form was used to obtain the data from hospital information system, and the two groups (two researchers per group) cross-checked one by one. Gender, age, all α-HBDH test results, exact exposure date (accurate to one day), disease onset date, date of transforming to severe and critical type, outcome, death date, etc. were collected. The data within the course of 30 days after symptom onset and 40 days after exposure onset were analyzed. The distributions of α-HBDH median value in course/period were plotted with an interval of 1 day, 2 days, and 5 days (T1, T2, T3…Tn represented the 5 days unit successively). In this cohort, symptom onset was regarded as disease onset and the corresponding date was set as the 1st day to record data from symptom onset. For patients with clear exposure date, the exposure date was set as the 1st day to record data when studying the α-HBDH changes from exposure onset. The day when the clinical type changed to severe or critical type was regarded as the “day 0” to study the distribution of α-HBDH before and after the type transformation. The death date was set as “day −1” and selected the −16th day to −1st day as the duration to calculate the α-HBDH median value per two days and plot the distribution when studying the α-HBDH features before death. Two respiratory physicians classified the patients from mild to critical type and then cross-checked, a third expert was involved when there was disagreement.

All patients’ general information, days from symptom onset to admission to hospital, from symptom onset to transformation to severe/critical type, from severe type to critical type, from symptom onset to death, from symptom onset to discharge, from exposure onset to discharge were included. The α-HBDH median value, abnormal percentage, and distribution of α-HBDH median over time were studied in different course with various initial time, and different periods, including after disease onset, exposure onset, 4 days before and after transformation to severe and critical type, and before death.

### 2.3 Statistical Analysis

All statistical analyses were performed through SPSS 20.0. Binary data were described using frequency and percentage. The normality of continuous data was checked. Mean and standard deviation were used to describe variables with normal distribution; otherwise, median (interquartile, IQR) was used. Categorical data were described as frequency (%); the chi-square test was applied to assess significance between groups. All graphs were processed by GraphPad Prism 8.0.

## 3 Results

### 3.1 General Information

Up to February 28, 542 patients with suspected and confirmed COVID-19 have been admitted to our hospital. A total of 142 cases were positive for nucleic acid test, among which the data of 9 cases were incomplete, 2 cases were infants, thus 131 cases were included in this study finally. 121 cases discharged from the hospital while 10 cases died. There were 63 males and 68 females, and the average age was 50.23±17.16 years old. The time from exposure to symptom onset was 11.32±8.48 days, symptom onset to hospital admission was 4.55±3.11 days, average hospitalization time was 26.92±9.25 days, symptom onset to severe type was 8.09±4.42 days, symptom onset to critical type was 11.58±5.79 days, time from severe to critical type was 3.87±4.78 days, symptom onset to death was 18.40±9.77 days, and symptom onset to discharge from the hospital was 25.77±10.61 days.

### 3.2 α-HBDH Results

The 131 cases underwent 37 laboratory indicators tests in outpatient and hospitalization. This study selected 565 tests of α-HBDH, accounting for 2.30% of the all results of the indicators. The normal range of α-HBDH value is 72-182 U/L.

### 3.3 The features of α-HBDH during the course from symptom onset

During the first 30 days, there were 506 tests and the α-HBDH median value was 156.33 (125.12-221.59) U/L, and the abnormal percentage was 35.97%. In T1-T6, the abnormal percentage of α-HBDH value was 26.87%, 43.86%, 40.00%, 34.38%, 35.06%, 29.03%, respectively. Among the changes in α-HBDH per day, α-HBDH median value increased from the 1st to the 4th day while decreased during the 15th to the 18th day, and fluctuated around upper limits of normal (ULN) from the 4th to the 15th day. And the peak value 191.14 (149.41-223.65) U/L appeared on the 8th day. After the 18th day, α-HBDH median value remained in the normal range, and the outliers on the 21st and the 27th day might related to some severe or critical cases. According to the change trend per 5 days, the α-HBDH median value had an increasing trend from T1 to T2, and a decreasing trend from T2 to T6. The peak 174.69 U/L appeared on T2. The α-HBDH median value was in the normal range during T1-T6.(Table1,Figure1)

**Figure 1.**
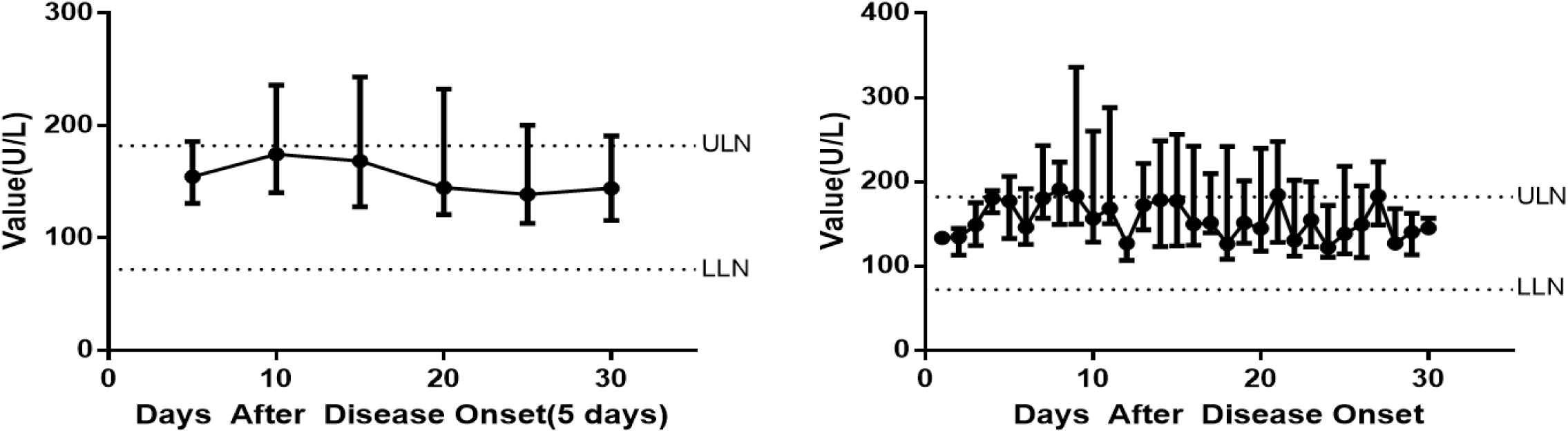
Changes of α-HBDH during the total course

**Table 1.** α-HBDH median and abnormal value in 131patients since symptom onset

| Period | Median value(U/L) | Total tests | Abnormal tests | Abnormal percentage |
| --- | --- | --- | --- | --- |
| First 30 days | 156.33(124.87-219.05) | 506 | 182 | 35.97% |
| T1 | 154.55(130.92-185.97) | 67 | 18 | 26.87% |
| T2 | 174.69(140.49-236.13) | 114 | 50 | 43.86% |
| T3 | 168.53(127.84-243.26) | 90 | 36 | 40.00% |
| T4 | 144.94(120.92-232.52) | 96 | 33 | 34.38% |
| T5 | 138.76(113.17-200.49) | 77 | 27 | 35.06% |
| T6 | 144.06(115.74-190.96) | 62 | 18 | 29.03% |

### 3.4 The features of α-HBDH in 42 cases during the course from exposure onset

We followed up all of the cases and finally defined the 42 cases with a exact exposure date. 42 cases underwent 206 tests, and the α-HBDH median value was 162.59 (131.91-225.31) U/L, the abnormal percentage was 42.72%. The changes of α-HBDH median value indicated the first test of α-HBDH since exposure onset appeared on 8th day, with the corresponding results as 180.28 (171.12-184.39) U/L, and it reached its peak on the 18th day. α-HBDH median value increased from the 8th day to 18th day, decreased from the 18th day to 24th day, and fluctuated within the normal range after the 24th day, the abnormal range was from the 10th day to 22nd day.(Figure2)

**Figure 2.**
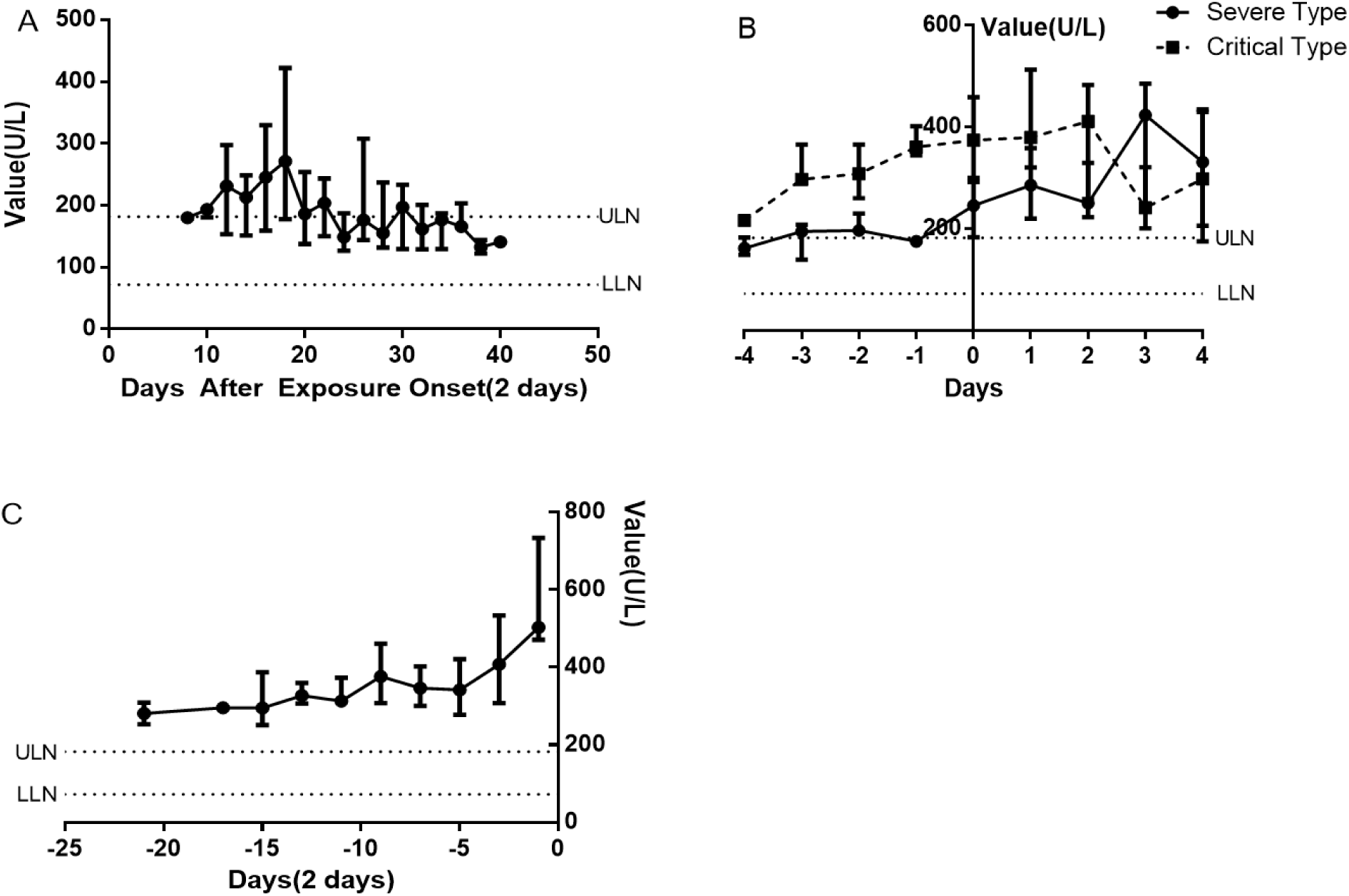
(A) Changes of α-HBDH since exposure onset, (B) Changes of α-HBDH before and after transforming to severe or critical type, (C) Changes of α-HBDH before death.

### 3.5 The features of α-HBDH during the period before and after transforming to severe and critical type

The date of transforming to severe type was set as “day 0” in 19 severe cases and 18 critical cases, and the changes during 4 days before and after “day 0” was plotted. In the 73 α-HBDH test results, the median value was 235.83 (175.37-331.44) U/L, and the abnormal percentage was 72.60%. The tendency showed that it just slight changed during −4th day (161.95 U/L) to −1st day (175.28 U/L). Among the period, α-HBDH median value was in the abnormal range except on the −1st day and −4th day. Similarly, the change trend during 4 days before and after transforming to critical type from 18 cases was plotted. The chart showed that the α-HBDH median value was increased from −4th day (216.49 U/L) to +2nd day (411.03 U/L). And all results situated in the abnormal range during the −4th day to +4th day.(Figure2)

### 3.6 The features of α-HBDH during the course before death

The α-HBDH median value was 332.50 (292.81-474.85) U/L during 1-21 days before death, and the abnormal percentage was 96.23%. α-HBDH median value fluctuated around 320U/L during the −21st to −5th day, nearly presenting a plateau level. However, α-HBDH median value increased sharply during the −5th to the −1st day, and all results were abnormal values from the −21st day to −1st day. (Figure2)

## 4 Discussion

According to the α-HBDH median value and the abnormal percentage in each time unit, it showed that abnormal results mainly appeared during T2-T4. The abnormal percentage of α-HBDH value in T2 was the highest, which was 43.36%, and the peak 174.69 (140.49-236.13) U/L also appeared in T2. It indicated that tissues and organs damage mainly occurred in first 10 days. α-HBDH median value showed an upward tendency during T1 to T2, and then gradually decreased, but the results was all in the normal range during T1-T6, which might be the effect of the weak increase of α-HBDH in most mild and moderate patients on the overall data.

As shown in the changes per day, the α-HBDH median value increased during the 1st day to 4th day while decreased during the 8th to 12th day and the 15th to 18th day, and it was basically in the normal range after the 16th day. The peak value 191.14 U/L was on the 8th day, abnormal tests were distributed within the 7th to the 9th day and the 13th to the 15th day, the outliers of the 21st day and the 27th day might be related to some severe and critical patients. The α-HBDH median value closed to the ULN on the 4th day since disease course, while the average time from symptom onset to admitted was 4.55±3.11 days, indicating that α-HBDH might have increased 1-2 days before admission. So there might be potential value for diagnosis in the early stage.

The time of symptom onset was clear and always regarded as the start of the disease, while the tissue might suffered damage before symptom onset because of the infected virus. Symptom occurs just when the viral load and damage reached a certain extent, thus the total course should be recorded starting from the day of exposure. After carefully follow-up, it was confirmed that the exposure time of 42 patients in our study could be accurate to a day, and the time from exposure to symptom onset was 11.32±8.48 days. In order to further study the changes of α-HBDH in the whole course including asymptomatic stage, we took the exposure onset as the first day, and performed a time change chart during 40 days after exposure onset. The first test of α-HBDH since exposure onset appeared on the day 8, with the results close to the ULN, indicating that the virus had caused tissue and organ damage before the symptoms onset. On the 18th day, α-HBDH median value reached the peak, indicating that tissue and organ damage accumulated to the maximum degree, and resulting in the highest level of α-HBDH. After the 24th day, α-HBDH median value gradually decreased to the normal range, which might be related to the protective effect of antibody.

Due to the differences in the length of incubation period and the progression of disease course, and there was also difference in the time for patients to transform from moderate type to severe type and critical type. In order to reflect the changes of α-HBDH during the transformation from moderate type to severe type and critical type more accurately, we observed and analyzed the changes of α-HBDH during 4 days before and after the transformation. According to the fact that the time for 18 critical patients change from severe to critical type was 3.87±4.78 days, we collected the results tested 4 days before and after the transformation date for statistical analysis. In the severe group, the α-HBDH median value fluctuated at the ULN from the −4th day to −1st day, and there was no obvious change. Moreover, α -HBDH median value rapidly increased and exceeded the ULN from the −1st day to the 4th day, indicating that the tissue and organ injury might be further aggravated after transforming to severe type. Similarly, α-HBDH median value increased from the −4th day to the 2nd day but decreased gradually after the 2nd day in the critical cases, and all results exceeded the ULN within the period, indicating a more serious injury of tissue and organ in this type.

The time from symptom onset to death was 18.30±9.65 days, which was relatively dispersed. Thus, it was difficult to describe the features of α-HBDH before death. In order to study the feature of change in this stage, we performed a time change chart of α-HBDH with the death date as “day −1”. The results showed that the α-HBDH median value fluctuated around 320 U/L during the −21st to −5th day, and there was no obvious change. α-HBDH median value increased rapidly 5 days before death, which indicated there was a persistent and irreversible tissue and organ injury before death, Furthermore, aggravated of injury happened within 5 days before death, and α-HBDH value over 320 U/L suggest a high risk of death.

α-HBDH distributes in many tissues and organs, mainly in the heart. In clinical work, we usually regard α-HBDH as a kind of myocardial enzyme. The increase of α-HBDH in severe type, critical type and death patients in this cohort indicates there might have the serious tissue and organ damage in these patients, and the higher the α-HBDH, the more serious the illness, and the worse the outcome.

However, this study has several limitations. All the patients in the study come from the single hospital, and the sample size is small. And we may fail to detect the daily α-HBDH value in patients and lose some information.

## 5 Conclusion

α-HBDH value increases in some COVID-19 patients, obviously in severe type, critical type and death patients, and mainly in 18 days after exposure onset and 10 days after symptom onset. α-HBDH increases 1 day before transforming to severe type, continues to increase in critical type and death patients, increases rapidly 5 days before death. The increase of α-HBDH suggests that COVID-19 patients have tissues and organs damage, mainly in heart. In brief, α-HBDH is an important indicator to judge the severity and prognosis of COVID-19 patients.

## Data Availability

Anyone who wishes to obtain the original data of this study with reasonable purposes can contact the correspondent author via email.

## Acknowledgements

We thank all colleagues who helped us during the current study. We thank all the medical staff involved in treating the patients and all patients and their families involved in this study.

## Author contributions

G.J.Q, M.L.Z and G.X.H performed the data collection. H.M.Z and B.P prepared the first draft of the manuscript, validated the data collection, performed the data analysis and edited the manuscript, G.J.Q, H.Y, G.X.H, L.C, S.S.W validated the data collection, edited the manuscript. B.P is the guarantor of the manuscript.

## Ethical standards

The ethics committee of the Xiangyang No1 Peoples’ Hospital Affiliated of Hubei University of Medicine approved this study, and granted a waiver of consent from the study participants (No. 2020GCP012)

## Data available statement

The data that support the findings of this study are available from the corresponding author, BP, upon reasonable request.

## References

1. Huang CL, Wang YM, Li XW, et al. Clinical features of patients infected with 2019 novel coronavirus in Wuhan, China. Lancet, 2020; 395 (10223): 497–506. doi:10.1016/S0140-6736(20)30183-5.

2. Chen NS, Zhou M, Dong X, et al. Epidemiological and clinical characteristics of 99 cases of 2019 novel coronavirus pneumonia in Wuhan, China: a descriptive study. Lancet, 2020; 395 (10223): 507–13. doi:10.1016/S0140-6736(20)30211-7

3. Alexandra LP, Rebecca K, Lawrence OG. The Novel Coronavirus Originating in Wuhan, China: Challenges for Global Health Governance. JAMA, 2020. doi:10.1001/jama.2020.1097

4. Li Q, Guan XH, Wu P, et al. Early Transmission Dynamics in Wuhan, China, of Novel Coronavirus-Infected Pneumonia. The New England journal of medicine, 2020; 382 (13): 1199–1207. doi:10.1056/NEJMoa2001316

5. World Health Organization. Novel Coronavirus (2019-nCoV) Situation Report-11. Available at:https://www.who.int/docs/default-source/coronaviruse/situation-reports/20200131-sitrep-11-ncov.pdf?sfvrsn=de7c0f7_4

6. World Health Organization. Coronarirus disease (COVID-19) Weekly Epidemiological Update. Available at:https://www.who.int/docs/default-source/coronaviruse/situation-reports/20200831-weekly-epi-update-3.pdf?sfvrsn=d7032a2a_4

7. Patrick O, Laurent J, Olivier M, et al. Prognostic value of C-reactive protein and cardiac troponin I in primary percutaneous interventions for ST-elevation myocardial infarction. AM HEART J, 2006; 152 (6): 1161–1167. doi:10.1016/j.ahj.2006.07.016

8. Dissmann R, Linderer T, Schröder R. Estimation of enzymatic infarct size: direct comparison of the marker enzymes creatine kinase and alpha-hydroxybutyrate dehydrogenase. Am Heart J. 1998;135(1):1–9. doi:10.1016/s0002-8703(98)70335-7

9. Kemp M, Donovan J, Higham H, Hooper J. Biochemical markers of myocardial injury. Br J Anaesth. 2004;93(1):63–73. doi:10.1093/bja/aeh148

10. Gungor K, Philip L, Abdul BK. Serum alpha-hydroxybutyrate dehydrogenase levels in children with sickle cell disease. The American journal of pediatric hematology/oncology, 1981; 3 (2): 169–171. doi:10.1097/00043426-198100320-00010

11. Apostolov I, Minkov N, Koycheva M, et al. Acute changes of serum markers for tissue damage after ESWL of kidney stones. Int Urol Nephrol. 1991;23(3):215–220. doi:10.1007/BF02550414.

12. Cen Y, Chen X, Shen Y, et al. Risk factors for disease progression in patients with mild to moderate coronavirus disease 2019-a multi-centre observational study. Clin. Microbiol. Infect, 2020; 26(9):1242–1247 doi:10.1016/j.cmi.2020.05.041

13. Zhao DH, Yao FF, Wang LL, et al. A Comparative Study on the Clinical Features of Coronavirus 2019 (COVID-19) Pneumonia With Other Pneumonias. Clinical infectious diseases, 2020; 71 (15): 756–761. doi:10.1093/cid/ciaa247

14. Dong YL, Zhou HF, Li MY, et al. A novel simple scoring model for predicting severity of patients with SARS-CoV-2 infection. TRANSBOUND EMERG DIS, 2020. doi:10.1111/tbed.13651

15. Li MY, Dong YL, Wang HJ, et al. Cardiovascular disease potentially contributes to the progression and poor prognosis of COVID-19.. Nutrition, metabolism, and cardiovascular diseases. NMCD 2020; 307(7): 1061–1067. doi:10.1016/j.numecd.2020.04.013

16. Zhang H, Liao YS, Gong J, et al. Clinical characteristics of coronavirus disease (COVID-19) patients with gastrointestinal symptoms: A report of 164 cases. Dig Liver Dis, 2020. doi:10.1016/j.dld.2020.04.034

17. Nationan Health Commissiom of the People’s Republic of China. Diagnosis and treatment of corona virous disease-19 (8th trail edition), 2020. Available at:http://www.nhc.gov.cn/yzygj/s7653p/202008/0a7bdf12bd4b46e5bd28ca7f9a7f5e5a/files/a449a3e2e2c94d9a856d5faea2ff0f94.pdf

